# Primary care physician and nurse practitioner perspectives on digital self-triage patient navigator tools: Lessons from Ontario for future design and acceptance

**DOI:** 10.64898/2025.12.09.25341895

**Authors:** Laurel C. Austin, Marisa L. Kfrerer, Cathy Faulds, Robert D. Austin, Christina Ziebart, Daniel Pepe

**Affiliations:** Ivey Business School, Western University, London ON; Department of Family Medicine, Schulich School of Medicine & Dentistry, Western University, London ON

**Keywords:** triage, patient navigation, primary health care, primary care physicians, self- assessment, medical informatics

## Abstract

**Aim:** To explore Ontario primary care physician and nurse practitioner experiences with, and perceptions of, digital self-triage patient navigator tools, and to identify implications for future design and implementation.

**Background:** Purported benefits of digital self-triage tools include triage any time/place, easily updated triage logic, and reduced primary care demand. The COVID-19 pandemic provided a particularly important context for studying digital self-triage; primary care clinicians are an important group to learn from, but their perspectives have received little attention.

**Methods:** We interviewed fourteen Ontario family physicians/nurse practitioners in Spring 2021 using semi-structured interviews. We asked about experiences with, and perceptions of, digital self-triage tools, including the province’s centralized tool. Interview data were analyzed using reflexive thematic analysis.

**Findings:** Participants had limited awareness of the provincial tool. They had greater awareness of locally developed tools that were intended to mimic the provincial tool’s logic, but that were integrated into primary care. Respondents noted local tools were embedded in EMR systems, enabled scheduling, and were designed by others they knew and trusted. Respondents preferred self-assessment tools positioned as adjuncts to, rather than replacements for, clinical care, due to concerns about coordination and continuity of care. Concerns centred on tool efficacy, user input and interpretation, and access and equity.

**Conclusions:** This study highlights a gap between stand-alone digital self-triage tools (meant to reduce demand on primary care) and primary care. Locally developed tools intended to mimic that centralized tool, but integrated into primary care, are preferred. This aids primary care coordination and workflows but undermines ability to provide standardized triage anywhere, at any time, via a centralized tool, key benefits of digital self-triage. Future self-triage tools may be more effective if centrally maintained, but interoperable with local systems, developed with physician input, and customizable to support clinical workflows and continuity of care.

## Introduction

Digital self-triage is intended to help individuals assess themselves and receive advice on whether to seek care, where to seek it, and with what urgency (Semigran *et al*., 2015). Benefits include access to triage anywhere at any time, increased system efficiency, reduced demand on physicians, and reduced patient and physician exposure to communicable diseases (Jensen *et al*., 2020; Judson, Odisho, Neinstein, *et al*., 2020; Kellermann *et al*., 2010; Naved and Luo, 2024). With tools centrally maintained, software updates allow new knowledge to be efficiently and consistently incorporated into risk assessment, offering advantages over human systems where cognitive and organizational factors might impede such incorporation (Naved and Luo, 2024).

Such tools can also collect data to predict healthcare demand (Denis *et al*., 2021; Naved and Luo, 2024; Runkle *et al*., 2021).

Patient navigators are meant to eliminate barriers to timely health care by providing patient education and facilitating or coordinating medical care (Freeman, 2012; Peart *et al*., 2018). Traditionally a role played by humans, there is growing interest in digitizing patient navigator functionality (Offodile *et al*., 2021; Seitz *et al*., 2025). During the COVID-19 pandemic, timely access to triage, information, and care was a great concern in overwhelmed health care systems.

In response, jurisdictions and health care systems around the world rapidly deployed digital COVID-19 self-triage tools (Ziebart *et al*., 2023). Some of these incorporated patient navigator functionality, for example, providing educational information (Denis *et al*., 2021; Galmiche *et al*., 2020) and pathways to physicians and testing (El-Wahab and Metwally, 2021; Jensen *et al*., 2020; Judson, Odisho, Young, *et al*., 2020; Runkle *et al*., 2021). We refer to digital self-assessment tools that provide assessment, triage, and patient navigator functionality as digital self-triage patient navigator tools. We note that different self-assessment tools may have the same ‘look and feel’ but differ in important ways with respect to purpose and functionality.

Although the potential benefits of self-triage extend beyond infectious disease emergencies, the COVID-19 pandemic provided a particularly important context for studying digital self-triage because local, provincial, state, and national governments deployed centralized tools rapidly and at scale in order to manage demand and provide standardized triage and public guidance. These tools were often designed as a stand-alone application within a health care system, although a few were integrated into existing electronic health record systems (Judson, Odisho, Neinstein, *et al*., 2020).

In Ontario, Canada’s largest and most populated province, a centrally deployed provincial self-triage patient navigator tool was introduced in March 2020. The tool assessed COVID risk using if-then logic and provided advice based on symptoms, travel, and possible exposure. It incorporated relevant federal and provincial COVID-19 requirements and policies, advised on level of care required (e.g., go to an emergency department, get tested, self-isolate, watchful waiting), and provided patient navigation functions in terms of educational information, and, eventually, pathways to testing. It was reportedly visited more than one million times in its first week (Donovan, 2020) and was updated frequently as understanding about the virus evolved and testing became more widely available. At the same time, Ontario physicians and patients routinely encountered COVID-19 symptom checkers used for mandatory screening prior to entrance into health care settings, schools, daycares, and workplaces (Ontario Office of the Premier, 2020; Skrzypinski, 2020). These tools asked the same or similar questions. Thus, physicians were practicing within a broader digital ecology where multiple self-assessment tools could appear similar but differ in purpose and design.

Ontario’s provincial self-triage patient navigator tool formed part of the broader pandemic response. It was a stand-alone public-facing tool not integrated into primary care electronic medical record (EMR) systems. (In Ontario, there is not one common EMR system, but rather each medical facility can adopt one of several that are commercially available). Given that a goal of such tools is to reduce pandemic burden on primary care, primary care physicians are an important group from whom to learn. Their perspective can help illuminate what worked well, and what could be improved for use of digital self-triage, possibly including patient navigator functionality, in future emergencies such as pandemics, but also for self-triage with patient navigation in primary care more broadly.

## Research question

In this study we ask: What can we learn from Ontario primary care physicians’ and nurse practitioners’ experiences with, and perceptions of COVID-19 digital self-assessment tools, including the provincial self-triage patient navigator tool, in order to inform design and implementation of digital self-triage and patient navigation in future pandemics, and in primary care more generally?

## Methodology

### Study design

We conducted a qualitative descriptive study to explore Ontario primary care physicians’ perceptions of COVID-19 digital self-triage tools and use of self-triage in primary care more generally. Qualitative description is particularly suited to health services research where the goal is to provide a comprehensive, straightforward summary of participants’ views and experiences in their own words (Bradshaw *et al*., 2017; Sandelowski, 2000). Individual interviews with 14 primary care physicians or nurse practitioners were conducted (we refer to these as physicians going forward). Our research took place more than a year into the pandemic, when case counts were very high, province-wide stay-at-home orders were in effect, and the health care system was under extreme pressures. To ensure rigor in methods and reporting we followed the consolidated criteria for reporting qualitative studies guidance (COREQ) (Tong *et al*., 2007).

This study was approved by the [university name] Research Ethics Board.

### Participant recruitment

Participants were recruited between March and May 2021 via a professional newsletter, Twitter posts by physician researchers, targeted Facebook advertisements, and asking participants to share a study advertisement with colleagues. Materials stated we wished to talk with primary care practitioners “about your and your patients’ experiences with a more ‘digital-first’ approach to care during the pandemic.” Twenty-one Qualtrics-based pre-screening surveys were submitted, indicating interest in participating. All responses met the inclusion criteria of being a practicing Ontario primary care physician or nurse practitioner. (Exclusion criteria were not currently practicing primary care in Ontario). Each was invited by email to participate in an interview; fourteen accepted and were interviewed. A few participants were known by one or both physicians on our team (redacted initials); neither physician participated in conducting those interviews. Participants received a $40 retail gift card as a token of our appreciation. Our sample may be considered a convenience sample, possibly subject to selection bias. This may limit the transferability of findings.

### Data collection

Interviews were conducted over Zoom between April-May 2021 (Zoom Video Communications Inc., 2016). Two researchers experienced in conducting in-depth interviews participated in each interview (from among initials of 4 researchers redacted). Following verbal consent, interviews were recorded, transcribed, and then corrected/verified by a researcher. Transcripts were not reviewed by participants, due to pandemic burden on them at the time of data collection.

A semi-structured interview guide explored experiences with digital-first approaches to care during the pandemic. This analysis focuses on discussion about experiences with self-assessment tools generally and the provincial self-triage patient navigator tool specifically (recognizing physicians may have encountered multiple self-triage tools), whether the participant pointed patients to any self-assessment tools, perceived trustworthiness of user responses, and thoughts about digital self-triage more generally in primary care. Other parts of the interview asked about digital-first virtual care during the pandemic, to be reported elsewhere.

### Positionality

The research team included two Ontario primary care physicians, one female and one male (initials redacted), whose expertise in primary care and pandemic policies and practice informed interpretation of context-rich data. Two researchers were social scientists with extensive experience in qualitative research, one male professor in digital innovation (initials redacted), and one female professor of behavioural decision theory (initials redacted). Two were female PhD students in rehabilitation health science and with clinical experience (initials redacted). Together these brought varied expertise relevant to interpretation of the verbal data we’d collected. Three (initials redacted) had helped develop a digital self-triage tool for a local provincial health unit early in the pandemic, bringing expertise in this type of tool.

### Data analysis

We employed reflexive thematic analysis, as outlined by Braun and Clarke (Braun and Clarke, 2019, 2021; Clarke and Braun, 2017). This approach allowed us to focus on identifying and interpreting patterns in the data, taking a flexible, iterative, and reflective approach to inductive coding and generating themes. We followed Braun and Clark’s six phases recursively. Analysis began with familiarization as data were collected. Then each of the six co-authors independently read and a subset of interview transcripts to immerse themselves in the content and identify initial codes and themes. At least two researchers read each transcript, allowing different perspectives to be brought to data interpretation during frequent reflexive discussions.

Transcripts were re-read and codes were attached to specific segments of data, capturing meaningful elements of participants’ beliefs and experiences. More thorough coding was completed using MaxQDA24 software (VERBI Software, 2024), allowing us to systematically capture and review data relevant to proposed themes. Participants did not provide feedback on findings.

## Results

### Participants

Twelve physicians and two nurse practitioners were interviewed (n = 14; nine female, five male). Years in practice varied between one to twenty-three years. Most described patient populations as ‘cradle to grave’. Participants served in rural areas, semi-rural towns, and small and large urban areas, including Toronto. Many provided care in multiple settings (clinic, ED, nursing home, etc.). Three practiced in Indigenous communities, one exclusively. Two held university faculty appointments. Interviews lasted 28-49 minutes, averaging 36 minutes.

### Themes

We generated three main themes from the data, two of which have subthemes (see Table 1). The main themes are: 1) Participants had limited awareness of the provincial self-triage patient navigator tool but greater awareness of locally developed tools; 2) Participants were selectively open to digital self-assessment in primary care; and, 3) Concerns about digital self-assessment tools. We discuss each of these in turn.

**Table 1:** Themes identified in interview data.

| Main Themes | Subthemes |
| --- | --- |
| 1. Respondents had limited awareness of the provincial tool but greater awareness of locally developed tools | 1.1 Limited awareness of the provincial tool<br>1.2 Greater awareness of locally developed tools |
| 2. Respondents were selectively open to digital self-assessment in primary care |  |
| 3. Concerns about digital self-assessment tools | 3.1 Concerns about tool accuracy and effectiveness<br>3.2 Concerns about user input and interpretation<br>3.3 Concerns about access and equity |

## Theme 1: Participants had limited awareness of the provincial tool but greater awareness of locally developed tools

### 1.1 Limited awareness of the provincial tool

A dominant finding was limited awareness and use of the provincial tool. Most participants (n=11) said they were not very familiar with it, were unsure if they had ever used it, or reported having never seen it. For example, one vaguely recalled “possible” use as a “prelude to a COVID test” for self or family members (P12). P12 continued, “*Most [patients] don’t seem to be aware of the tool. It doesn’t come up in conversation [with patients]…. to be frank, a lot of doctors are confused about where to point patients.*”

A few did report pointing patients to the provincial tool, using it in their clinic as a screening tool for providers or patients, or using it together with patients. While awareness of the provincial tool was low among respondents, one noted, *“It probably is reducing the amount of phone calls we’re getting; it’s hard to know exact numbers …. but at least it gives people another option for trying to find that information*” (P07).

### 1.2. Greater awareness of locally developed tools

Respondents were more likely to discuss pointing patients to locally developed tools than to the provincial tool. P05 pointed patients to a local hospital website that, “*had a really nice pathway to getting you into a booking.*” Two mentioned pointing patients to “*the Toronto tool*” developed by a local physician. Others pointed patients to local public health unit self-triage tools, some of which offered more patient navigator functionality than the provincial tool. For example, one described a tool that interfaced directly with care workflows: “*Local physicians had brought back to me that they would prefer to have a local option with local physicians doing it. One of the benefits is that you know who’s seeing the patients…we set up a virtual walk-in clinic*” (P04).

Some had integrated digital COVID-19 self-assessment tools into their clinical practice, often mentioning that provincial resources including the provincial tool informed questions they asked. These included digital forms in EMR systems, sent before an appointment, or tablets provided in clinic pre-visit. P01 explained, “*having the results be directly imported into the EMR is crucial*”. Some had personally participated in creating these tools while others relied on office managers or other physicians. One described having changed a question found on the provincial tool because they felt the advice given was not appropriate.

## Theme 2: Participants were selectively open to digital self-assessment in primary care

Participants expressed mixed views about future digital self-triage with patient navigator functionality in primary care. Some were open to the idea of tools that independently triaged and directed patients to care. Others were not. However, some who were skeptical of self-triage tools that would navigate to care were open to digital self-assessment tools that collect information pre-visit to support clinical workflows and open time for patient care.

Those open to the idea of digital self-triage tools more generally all placed some caveats on this acceptance. One noted specific conditions for which they felt this would be appropriate, including rashes and bladder infections. Another explained such tools must share information with primary care physicians, otherwise continuity of care would suffer. P05 thought the idea of self-triage could be beneficial as long as it was not the only venue to triage:

> “*I think it has value. I think it’s very dangerous for that to be the single pathway. I certainly believe there should be multiple channels available…I’m very interested in these chat bots with algorithms for this sort of thing*.”

Many of the participants were not open to substitutes for triage and navigation that traditionally has been their responsibility. Using COVID-19 tools as an example, P02 stated, “*if the purpose of the tool is to tell you go get an assessment or get a COVID swab then sure, yeah, they’re fine. But if they are to be a surrogate for a physician assessment, then I’d say no*.”

However, some in the group skeptical of self-triage more generally were open to digital self- assessment tools that would collect data pre-visit to improve clinic workflows and inform their own care provision. Several said that such tools would only be of use if integrated into EMR systems. Some shared they already had such tools in their clinic: “*I do use those types of tools already for people with abdominal pain, people with chest pain, pre-natal… they’ve already written it all down 24 hours before. My note is half done because it input straight into the EMR… I can just spend more time with the patient*” (P02).

Other respondents suggested that future self-assessment tools must be evidence based, developed with currently practicing physicians, and backed by trusted medical organizations in order to build physician and patient trust and uptake.

## Theme 3: Concerns about digital self-assessment tools

Participants shared several concerns that led them to be cautious in their embrace of digital self-assessment tools. These concerns can be grouped into concerns about: 3.1) Tool accuracy and effectiveness; 3.2) User input and interpretation, and 3.3) Access and equity.

### 3.1 Concerns about tool accuracy and effectiveness

We heard several concerns related to self-assessment tool performance. Some participants were concerned that there has been little study of digital self-assessment tool efficacy, including Ontario’s self-triage tool. One noted that because the provincial tool had not captured identifying information, there was no way to track and know if use had reduced unnecessary trips to emergency rooms. Others suggested over-diagnosis could be a concern: “*I think ultimately services like that, from a medical, legal perspective, are going to recommend that most people seek assessment, one way or the other. So, I don’t know if it necessarily saves any time*” (P06).

Several felt that asymptomatic/mildly symptomatic cases would be missed more often by digital tools than by humans due to inability to ask follow-up, nuanced questions. Some of these concerns led to preferences for pre-clinical self-assessment tools rather than full-fledged self-triage tools.

### 3.2 Concerns about user input and interpretation

Beyond concerns about tool performance, participants also worried about how patients interpret and respond to digital self-assessment questions. We heard concerns that some patients don’t fully understand how to translate symptoms into the limited response options available on digital tools (e.g., “*do you say ‘yes’ if you’ve had a cough for two months*?”). Other participants felt that some patients who have already concluded they do (not) have a condition select responses consistent with that pre-conception: “*they kind of just lead themselves to the answer they want; if I actually talk to somebody I can go back and forth and ….[get] a little bit more information*” (P03). These concerns led some respondents to prefer pre-visit self-assessment tools, with triage completed by the provider.

### 3.3 Concerns about access and equity

Several participants highlighted the importance of equity and accessibility considerations when deploying digital self-assessment tools, highlighting the “*digital divide that exists in the Canadian population*” (P12). Some fretted that certain subgroups, e.g., elderly patients, patients without access to technology/internet, or those with limited finances and phone data limits might struggle with, lack access, or be unwilling to use digital tools. Ensuring that assessment tools are offered in various forms (i.e. computer-based, telephone, and in-person) or perhaps having in-clinic staff help with use was seen as important.

## Discussion

Our findings suggest that the value of a centralized digital self-triage tool to primary health care depends not only on the quality of its triage logic and accessibility to patients, but also on whether it can connect to local care delivery. Ontario’s provincial self-triage tool offered the potential advantages of centralized digital triage, including easy access from anywhere at any time, consistent guidance and -- relevant during a pandemic -- a single point at which rapidly changing scientific understanding of disease could be integrated into triage. However, perhaps because it was a stand-alone patient-facing tool that did not integrate into local EMRs or workflows, many respondents were more aware of, and more likely to use or point patients to, locally developed alternatives (including ones developed in-house) that offered better supports and navigation functionality, for example, access to scheduling or virtual care, and pre-visit assessment documented in the clinic’s EMR system. It seems unlikely that all hospitals and clinics had the resources to continuously update these tools as knowledge grew. Thus, these local tools may have proved useful in day-to-day primary care, but their proliferation may also have undermined the promised benefits of the ability to maintain one up-to-date triage logic and provide consistent advice across settings that come with one centrally maintained tool. Our findings therefore suggest that future centrally deployed self-triage tools, especially in time of public health emergencies, may offer additional benefits to primary care if they are designed for interoperability with local clinical systems and workflows.

Our data suggest that co-design of tools, including the internal logics, with practicing primary care physicians will be a key factor in building physician trust and adoption (Göttgens and Oertelt-Prigione, 2021; Scariot *et al*., 2012). A central tool that could be integrated into local systems would allow a central authority to more comprehensively collect data and predict care demand. Integration into local systems, with possibilities to incorporate the central tool and add functionality that meets local needs, such as scheduling, pre-visit data collection, etc., would provide additional benefits in primary care. Importantly, integration that enables family doctors to see patient use of the central tool would enhance continuity of care because physicians would know who had used the tool and what advice had been given.

With respect to digital self-triage tools that offer patient navigation in primary care more generally, while some of our respondents were open to this idea, more were comfortable with the idea of digital self-assessment tools that support their own clinical assessment, rather than self-triage tools that assess, triage, and provide navigation to care. The potential of such tools to reduce unnecessary phone calls and free physician’s time for care was highlighted. Several respondents already used such tools for other conditions, and research has found that pre-consultation history-taking systems can improve clinical efficiency, support diagnostic accuracy, improve patient-physician communication, and reduce documentation time (Wickramasekera *et al*., 2023; Zhakhina *et al*., 2023). Evidence from the UK suggests that such tools are also beneficial in systems with a dedicated triage-GP (Gupta *et al*., 2026).

While there has been little research in this domain, respondents’ concerns about assessment quality have been validated by research that shows, for example, that different COVID-19 self-assessment tools come to different conclusions, provide different advice, differ in probability of advising to seek care, and differ in performance compared to human physicians (Hammoud *et al*., 2024; Mansab *et al*., 2021). Our work suggests that acceptance of self-triage more generally may increase for tools that have been researched and found to not have such limitations, in addition to addressing physicians’ concerns about the quality of questions asked, patient ability to respond appropriately, and accessibility and equity. Our study suggests that having physicians involved in tool development is important for acceptance, and critically, such tools need to be designed to facilitate, not harm, coordination and continuity of care, two key functions in primary care (Jimenez *et al*., 2021; Starfield, 1998).

## Limitations

Our sample was small, a convenience sample, and limited to Ontario; thus, findings should be interpreted as exploratory and hypothesis-generating rather than definitive. Recruitment was conducted via social media and professional networks; respondents were only minimally compensated for their time via a small gift card. Any of these factors may have introduced unknown selection bias into our sample. This may limit the transferability of findings to other regions or health systems, especially those that differ in how COVID-19 self-triage patient navigators were designed and implemented. However, we did hear a range of beliefs within responses, suggesting this was not an overly homogeneous sample.

While our data were collected five years ago, COVID-era tools remain the largest ‘natural experiment’ in centralized digital self-triage; lessons learned are directly relevant to future pandemics. Further, our work applies to self-triage more generally, and there has been little study of physician perceptions of self-triage. It’s important to note that several of our respondents spoke of how overwhelming the pandemic had been for physicians and their staff. The fact they took time to talk to us suggests they took this research effort seriously; we feel it’s important to share what can be learned from the experiences and perceptions shared in the interviews.

## Conclusions

This study suggests that stand-alone centralized self-triage tools that are widely available to patients and meant to reduce demand on primary care providers may have limited salience in primary care. In fact, given a stand-alone self-triage tool that cannot connect to local EMRs or workflows, primary care organizations may respond by creating or relying on their own locally developed tools that are explicitly intended to mimic the centralized tool’s decision logic, but that are integrated into primary care practice. This preserves local usefulness, but undermines one of the main advantages of centralized digital triage: a single, centrally updated source of standardized triage logic and guidance. Future centralized self-triage tools, especially in pandemics and other public health emergencies, may therefore offer additional benefits if designed for interoperability with local clinical systems by allowing health systems to retain the benefits of centralized updating and more consistent triage while also supporting continuity of care and primary care workflows. At present, digital self-assessment tools appear most acceptable in primary care when used as adjuncts to clinical care rather than substitutes.

However, many primary care physicians are open to the idea, provided concerns about tool performance, usability, and equitable access are addressed by high-quality research.

## Declarations

## Data Availability

This project received ethics approval from Western University Health Science Research Ethics Board. The Letter of Information (LOI) that participants signed says no identifiable data will be shared outside the research team. Given in-depth interviews, the nature of our data does not allow us to completely disentangle possibly identifiable information from the dataset, so we cannot share that in a repository and abide by our LOI. Many de-identified excerpts from interviews are found within the manuscript. Researchers requesting access to the data more broadly should reach out to Western University Health Science Ethics Review Board at.

## Acknowledgements

We gratefully acknowledge Western University’s support of this work through an internal COVID-19 Catalyst Grant. We are grateful to Kelly Zhang Zheng for work as a research assistant on the project. We are especially indebted to the fourteen providers who took time during what was probably the busiest and most stressful time of their professional lives to participate in this research.

## Financial Support

This research was funded by Western University, Grant Number R5904A03. The funder had no role in design, data collection, analysis, writing or decisions about publication of this article. The funder didn’t influence the results/outcomes of the study despite author affiliations with the funder.

## Conflicts of Interest

None.

## Ethical Standards

The authors assert that all procedures contributing to this work comply with the ethical standards of the relevant national and institutional guidelines on human experimentation (Canada TCPS2 2022; Western University) and with the Helsinki Declaration of 1975, as revised in 2008.

Verbal informed consent was obtained from all subjects. Verbal consent was witnessed and formally recorded.

## Notes

### Competing Interest Statement

The authors have declared no competing interest.

### Author Declarations

Ethics approval was received from the University of Western Ontario Health Science Research Ethics Board, reference number 118202.

### Summary of Updates

The revised manuscript sharpens the paper's central findings, particularly around how providers feel about digital self-triage more generally, without changing the underlying data or sample (14 Ontario family physicians/NPs, Spring 2021 interviews). The title now explicitly names nurse practitioners as study participants, better reflecting the actual sample (12 physicians, 2 NPs) and broadening the framing from "family physicians" to "primary care" perspectives throughout. We restructured themes for clarity, especially on general attitudes toward self-triage: The most substantive change is in Theme 2/3. In the original version, "perceived value" was framed somewhat dichotomously across two subthemes (tools playing a positive public health role vs. being supportive-but-not-a-substitute). The revised version replaces this with a single, more nuanced theme (Participants were selectively open to digital self-assessment in primary care) that better captures the conditional, mixed nature of provider attitudes (e.g., openness to self-triage for specific conditions, interest in chatbot-assisted triage, but resistance to tools replacing physician judgment). This directly addresses the goal of clarifying how providers feel about self-triage as a general concept rather than presenting split findings. In addition, three concerns sub-themes were relabeled and reorganized for precision. The abstract and conclusions were revised to match the new theme structure and to state findings more directly (e.g., "digital self-assessment tools appear most acceptable in primary care when used as adjuncts to clinical care rather than substitutes"). Discussion was strengthened with new evidence: Several new citations were added (e.g., on symptom-checker performance variability, human-centered design, pre-consultation history-taking systems, UK triage-GP models) to better ground physicians' stated concerns and preferences in existing literature.

